# Retinal Vein Occlusion at the University of Gondar Tertiary Eye Care and Training Centre, North-West Ethiopia

**DOI:** 10.1101/2022.07.29.22278195

**Authors:** Matias Tilahun, Asamere Tsegaw

**Affiliations:** Debretabor University, Ethiopia; Department of Ophthalmology, College of Medicine and Health Sciences, University of Gondar, Ethiopia

**Keywords:** Retinal vein occlusions, Clinical pattern, Gondar, Ethiopia

## Abstract

**Objective:** Retinal vein occlusion (RVO) is the second most common retinal vascular diseases after diabetic retinopathy. Delay in detection and treatment can result in irreversible visual impairment and blindness. The aim of this study was to assess the magnitude and clinical pattern of patients with RVO presentedd to the retina clinic at University of Gondar Tertiary Eye Care and Training Centre

**Methods:** A hospital based Cross-sectional study was conducted from October 2017 – March 2018 and patients of all ages with RVO seen at our retina clinic during the study period were reviewed. Pertinent ophthalmic history, ophthalmic clinical examination and laboratory tests were done including detailed funducopy for each patient. Data were collected with structured questionnaire, entered to SPSS version 20 and analysed.

**Result:** A total of 38 eyes of 36 new patients with RVOs were seen during the six month study period and reviewed. Twenty four (66%) study patients were males and the mean age was 58 ± 10.87 years. Thirty four (94.4%) patients had unilateral disease. Nineteen (52.78%) had Central retinal vein occlusion (CRVO), 13 (36.11%) had branch retinal vein occlusion (BRVO) and 4 (11.11%) had hemispheric retinal vein occlusion (HRVO). Glaucoma was the commonest risk factor seen in 17 (47.22%) patients followed by systemic hypertension 10 (27.78%) and diabetes mellitus 8 (22%). The commonest complications encountered were macular edema, retinal or optic disc neovascularization and neovascular glaucoma seen in 15 (41.67%), 11 (30.5%) and 4 (11.11%) patients respectively. Over a third of patients 15 (41.67%) presented to our retina clinic after 6 months of onset of the illness and 15 (39.47%) eyes were blind at presentation.

**Conclusion:** Glaucoma, hypertension and diabetes mellitus were the most common risk factors identified among study patients. A majority of patients had potentially blinding complications. There was also delay in presentation. Diagnostic and therapeutic facilities of the center should be improved to prevent vision loss from complications. People should be educated to seek health care immediately after the onset of visual symptoms.

## INTRODUCTION

Retinal diseases are emerging causes of visual impairment and blindness in the world and retinal vein occlusion(RVO) is the second most common retinal vascular diseases after diabetic retinopathy. (1,2)

Ocular and systemic conditions like glaucoma, hypertension, diabetes mellitus and hypermetropia are found to be associated with increased risk of RVO.(3,4) Late detection and delayed treatment of RVOs can result in irreversible visual impairment and blindness from vision threatening sequelae(5)

Several studies conducted in the Caucasian population aged from 40 years and above have shown that the prevalence of branch retinal vein occlusion (BRVO) and central retinal vein occlusion (CRVO) ranges between 0.6% - 1.1% and 0.1–0.4%, respectively(6). In the few Asian studies, the prevalence of BRVO and CRVO was 0.6% and 0.2% in those more than 40 years of age. RVO was the most common retinal vascular disease in a population based study done in Nepal(5).

A population based study was done in central India to determine the prevalence and associations of RVO in 2013. A total of 4711 subjects were involved and the prevalence of RVO was 0.77%. RVO was significantly associated with older age, hypertension, narrow anterior chamber angle and higher blood urea concentration. (7)

A retrospective hospital based study done in the University of Port Harcourt Teaching Hospital, Nigeria showed that out of the 364 patients seen at the retina clinic during the study period, 27 (7.4%) had RVO. Systemic hypertension, diabetes mellitus, hyperlipidemia and glaucoma were the main risk factors identified. CRVO (74%) was more predominant than BRVO (26%). (8)

Another retrospective hospital based study done in a tertiary hospital in Nigeria in 2016 included 20 patients 14 (70.0%) males and 6 (30.0%) females with a mean age of 62.7 ± 10.4 years. Fifteen (68.2%) eyes had CRVO, 3 (13.6%) had BRVO, and 4 (18.2%) eyes had HRVO. Risk factors identified included hypertension in 14 (70.0%), diabetes mellitus in 9 (45.0%), and glaucoma in 5 (22.7%) of the study patients. Multiple risk factors were present in 14 (70.0%) patients. Complications included macula edema 15 (68.2%), retinal neovascularization 5 (22.7%), neovascular glaucoma 3 (13.6%), and vitreous hemorrhage 2 (9.1%). (2)

Available literature on the pattern and clinical profile of RVO is insufficient in this part of the world and there are only few studies done in Africa. So far, we do not have a study done on RVOs in Ethiopia. This study was, therefore, designed to add input on the scarce knowledge and fill the existing gap.

## Patients and Methods

### STUDY DESIGN AND PERIOD

A Hospital based prospective cross sectional study was conducted from October 2017 to March 2018 at the University of Gondar tertiary eye care and training center.

### STUDY AREA

The study was conducted at University of Gondar Tertiary Eye Care and Training Center, which is one of Ethiopia’s main eye care and training centers. It serves an estimated 14 million people living in North-West Ethiopia. The center provides eye care services both at base hospital and rural outreach sites. On average some 12,000 patients are seen at the retina clinic of the study center annually.

### STUDY POPULATION

All new patients with retinal vascular occlusion who visited the center during the study period were included in the study. Follow up cases, and those with ocular media opacities for funduscopy examination were excluded from the study.

### DATA COLLECTION PROCEDURE

Comprehensive Ophthalmic evaluation which included detailed demographic information, presenting complaint, associated ocular and systemic symptoms, past treatment history for ocular or systemic illnesses such as hypertension were taken. Presenting visual acuity; intraocular pressure measurement, slit-lamp examination of the anterior and posterior segment was performed.

Grading of visual impairment and blindness was done according to the WHO classification system as follows: visual acuity better or equal to 6/18 – normal; visual acuity between 6/24 and 6/60 – moderate visual impairment; visual acuity less than to 6/60 and better than or equal to counting fingers at 3m – severe visual impairment; visual acuity less than counting fingers at 3m – blindness; the results for the eye with the best presenting visual acuity were recorded.

Posterior segment evaluation was performed with +90D funduscopic lens on the slit-lamp (slit-lamp biomicroscopy with 90D lens) for each patient by a vitreoretinal specialist after dilation of the pupil with 1% tropicamide eye drop. Indirect ophthalmoscopy with +20D lens was used to examine selected patients. Pertinent and supportive laboratory investigations such as fasting blood sugar, serum lipid profile were also done for all patients.

OCT and fluorescein angiography studies were not done due to lack of these imaging modalities at the center during the study period.

### DATA ANALYSIS

The collected data was checked for accuracy, coded and entered in to SPSS Version 20 and analysed; P value of less than 0.05 was considered as statistically significant.

## Ethical Clearance

The study was conducted in full compliance with the 1964 Helsinki declaration on research involving human subjects. Prior to commencement of the study, ethical clearance was obtained from the Ethics Committee (Institutional Review Board) of University of Gondar.

## Results

We studied 36 (38 eyes) patients with RVO and the mean age of participants was 58 ± 10.87 years. This constituted 5% of patients with vitreo-retinal diseases seen during the study period. A Majority of participants 24 (66.6%) were males and 34 (94.4%) were Christians. Twenty one (58.33%) of them can’t read and write and 19 (52.78 %) were from rural areas. Farmers accounted for 18 (50%) of patients while 13 (36%) patients were government employees. The commonest age group was from 55-70 years 16(44%).

**Table I:** Socio-demographic characteristics of patients with RVO at Gondar University Hospital tertiary eye care and training center, North West Ethiopia, 2018 (n=36)

| **Variables** | **Category** | **Frequency/%** |
| --- | --- | --- |
| **Age group (years)** | 40-55 | 15 (41.67%) |
|  | 56-70 | 16 (44%) |
|  | >70 | 5 (13.89%) |
| **Sex** | Male | 24 (66.6%) |
|  | Female | 12 (33.33%) |
| **Occupation** | Farmer | 18 (50%) |
|  | Gov't employee | 13 (36.1%) |
|  | Other | 5(14%) |
| **Residence** | Rural | 19 (52.78%) |
|  | Urban | 17 (47.22%) |
| **Educational Status** | Can't read and write | 21 (58.33%) |
|  | High school | 1 (2.78%) |
|  | College and above | 14 (38.89%) |
| **Religion** | Christian | 34 (94.4%) |
|  | Muslim | 2 (5.56%) |

Reduction of vision was the commonest symptom of presentation which was seen in all patients with RVO. Six (16.67%) and 2 (5.56%) patients had metamorphopsia and eye pain respectively. Eight (22.22%) patients had relative afferent pupillary defect (RAPD). Only 11 (30.56 %) patients presented within a month of the onset of symptoms.

Fifteen (39.47%) eyes were blind, 9 (23.7%) had severe vision impairment, 14 (36.8%) had moderate vision impairment at presentation. Only 2 (5.56%) study patients had bilateral involvement.

**Table II.**
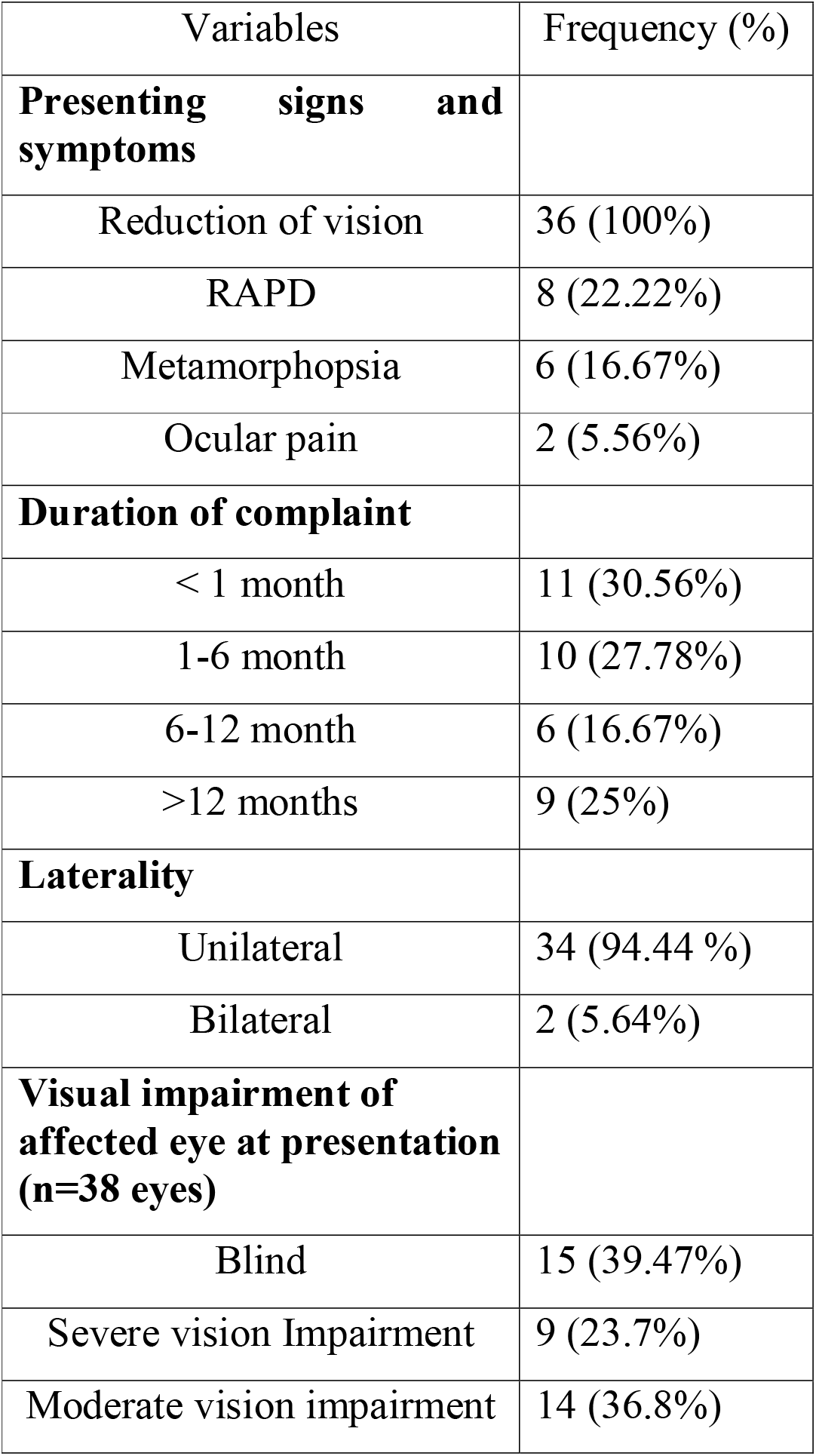
Clinical characteristics of patients with RVO in University of Gondar tertiary eye care and training center, 2018 (n=36)

In our study, Central Retinal Vein occlusion (CRVO) 19 (52.78%) was the commonest type of RVO followed by Branch Retinal Vein Occlusion (BRVO) 13(36.11%) and Hemispheric Retinal Vein Occlusion (HRVO) 4 (11.11%).

Blindness of the affected eye at presentation resulted from CRVO in 10 (26.3%) eyes and from BRVO in 5 (13%) eyes. CRVO was a cause of severe vision impairment in 6 (16%) of the study eyes and BRVO caused severe vision loss in only 2 (5%) of eyes. But there was no statistically significant association between the type of RVO and degree of visual impairment at the time of presentation (p=0.2315).

Macular edema 15 (39.47%) was the commonest complication identified in our study patients followed by retinal neovascularization 6 (15.8%) and disc neovascularization 5 (13.1%).

**Table III.** Complications in patients with Retinal vascular occlusion (RVO) presented to University of Gondar Hospital Tertiary Eye Care and Training center, North West Ethiopia; 2018 (n=38).

| **variables** | **Frequencies (%)** |
| --- | --- |
| Macular edema | 15 (39.47%) |
| Retinal neovascularization | 6 (15.8%) |
| Disc neovascularization | 5 (13.1%) |
| Neovascular glaucoma | 4 (10.52%) |
| Iris neovascularization | 2 (5.26%) |
| Vitreous hemorrhage | 2 (5.26%) |

Glaucoma 17 (47.22%) was the major risk factor identified followed by hypertension 10 (27.78%), hyperlipidemia 8 (22.22%), diabetes Mellitus 8 (22.22%) and hyperopia 3 (8.33%).

**Table IV:** Risk factors in patients with Retinal vein occlusion at University of Gondar Tertiary Eye Care and Training Center, North West Ethiopia; 2018 (n=36).

| **Variables** | **Frequency (%)** |
| --- | --- |
| Glaucoma | 17 (47.22%) |
| Hypertension | 10 (27.78%) |
| Diabetes mellitus | 8 (22.22%) |
| Hyperlipidemia | 8 (22.22%) |
| Hyperopia | 3 (8.33%) |

## Discussion

The incidence of retinal vein occlusion in our study was 5% which is relatively smaller than a hospital based retrospective study done in Nigeria which showed an incidence rate of 7.4%. (8) Population based studies in Australia, central India and Germany showed the prevalence of RVOs to be 1.6%, 0.77% and 0.4% respectively. This smaller prevalence was because of the fact that the studies were population based having a larger number of study participants.(7,9,10)

The commonest age range affected by RVO was 55-70 years with a mean age of 58 ±10 years. This is comparable with a study done in Nigeria 62.7 ± 10.4 years. Our study patients were younger compared to a study in Nepal (69.64 ± 7.31 years). (11) Males 24 (66.6%) were more affected by RVO and this is in agreement with a study done in Nigeria (70%) and a study done in Germany showed that men were 1.7 times more affected by RVO than females. (2,8)

In our study, CRVO (52.78%) was more common compared to BRVO (36.11%) and HRVO (11.11%). This is in agreement with a study done in a tertiary hospital in Nigeria in 2016 where CRVO (68.2 %) was the commonest followed by BRVO (13.6%) and HRVO (18.2%) (2). The results are also in agreement with another study in University of Port Harcourt Teaching Hospital, Nigeria where CRVO(74%) was more common than BRVO (24%). (8)

However, a hospital based study done in Nepal showed BRVO (70%) to be more common than CRVO (26.6%).(11) Other population based studies done in Korea, Australia and central India showed the prevalence of BRVO to be 63.4%, 69.5% and 85.7 % respectively.(1,7,10)

Atherosclerosis is thought to be a major cause of BRVO and patients from the developed countries are more prone to atherosclerosis compared to those from developing countries like ours because of their life style. In addition their study patients were older than ours. These might be reasons for BRVO to be more common in these studies.

Two (5.56 %) of our study patients had bilateral RVO and both resulted from CRVO. This is in agreement with a study done in a tertiary hospital in Nigeria in 2016 where bilateral disease was seen in 2 (10 %) of the study patients.(2) A population based study done in Australia showed bilateral disease in 3 (5.1%) of their study patients which is in agreement with our study. (7) Another study in University of Port Harcourt Teaching Hospital, Nigeria bilateral disease was seen in 7 (25.9%) of the study patients all of which were from CRVO. (12)

The commonest risk factors that were identified in our study were glaucoma 17 (47.22%), hypertension 10 (27.78%), diabetes 8 (22.22%), hyperlipidemia 8 (22.22%) and hyperopia 3 (8.33%). But they didn’t show a statistically significant association and this may be due to the small sample size in our study. These findings are also in agreement with a study done at University of Port Harcourt Teaching Hospital, Nigeria where their association was not statistically significant likely due to the small sample size. (12).

But these are the major statistically significantly associated risk factors identified in large population based studies done in Germany, Australia, Korea and central India.(1,7,8,11)

The major complications identified in our study were macular edema 15 (39.47%), retinal neovascularization 6 (15.8%), optic disc neovascularization 5 (13.1%), neovascular glaucoma 4 (10.52%), iris neovascularization (5.26%) and vitreous hemorrhage 2 (5.26%). These results are in agreement with a study done in a tertiary hospital in Nigeria in 2016 where macular edema (68.2%), retinal neovascularisation (22.7%), neovascular glaucoma (13.6%) and vitreous hemorrhage (9.1%) were among the major complications identified.(2)

The magnitude of macular edema was lower in our patients and this could be because of the fact that we were not using imaging modalities such as OCT and FA to make a diagnosis of macular edema and as a result some cases of mild macular edema might have been missed in our patients.

Fifteen (39.47%) eyes were blind, 9 (23.7%) had severe visual impairment, 14 (36.8%) had moderate visual impairment. CRVO was the major cause of blindness and severe visual impairment in 10 (71.2%) and 6 (66.67%) of the study patients respectively. But the difference was not statistically significant (p= 0.2315) with confidence interval of 95%. These findings are in agreement with a research done in University of Port Harcourt Teaching Hospital, Nigeria where 21 (61.7%) of blind eyes resulted from CRVO. (12) In a population based study done in Australia, 60 % and 14 % of the study eyes had best corrected VA of < 6/60 from CRVO and BRVO respectively and the association was statistically significant.(7)

## Conclusions

The findings of our study showed that the incidence of RVO was 5% and CRVO was the predominant type of RVO seen among study patients. Males were more frequently affected. Glaucoma, hypertension, diabetes, hyperlipidemia and hyperopia were the most frequently identified risk factors respectively. Macular edema, retinal and disc neovascularization and neovascular glaucoma were the top three complications detected. Fifteen 15 (39.47%) eyes were blind at presentation and 25 (69.44%) patients presented to the retina clinic 1 month after the onset of the illness.

Proper diagnostic and therapeutic set ups must be established to prevent vision loss from complications. People should be educated to seek health care immediately after the onset of symptoms.

## Data Availability

All relevant data are
within the manuscript and its Supporting
Information files.

## REFERENCES

1. Shin YU, Cho H, Kim JM, Bae K, Kang Mh, Shin JP, et al. Prevalence and associated factors of retinal vein occlusion in the Korean National Health and Nutritional Examination Survey, 2008–2012: A cross-sectional observational study. Medicine. 2016;95(44):e5185

2. Uhumwangho OM, Oronsaye D. Retinal Vein Occlusion in Benin City, Nigeria. Nigerian journal of surgery : official publication of the Nigerian Surgical Research Society. 2016;22(1):17–20.

3. Li J, Paulus YM, Shuai Y, Fang W, Liu Q, Yuan S. New Developments in the Classification, Pathogenesis, Risk Factors, Natural History, and Treatment of Branch Retinal Vein Occlusion. Journal of Ophthalmology. 2017;2017:4936924.

4. Hayreh SS, Zimmerman MB, Beri M, Podhajsky P. Intraocular pressure abnormalities associated with central and hemicentral retinal vein occlusion. Ophthalmology. 2004;111(1):133–41.

5. Thapa R, Bajimaya S, Paudyal G, Khanal S, Tan S, Thapa SS, et al. Prevalence, pattern and risk factors of retinal vein occlusion in an elderly population in Nepal: the Bhaktapur retina study. BMC ophthalmology. 2017;17(1):162.

6. Rogers S, McIntosh RL, Cheung N, Lim L, Wang JJ, Mitchell P, et al. The Prevalence of Retinal Vein Occlusion: Pooled Data from Population Studies from the United States, Europe, Asia, and Australia. Ophthalmology. 2010;117(2):313-9.e1.

7. Mitchell P, Smith W, Chang A. Prevalence and associations of retinal vein occlusion in australia: The blue mountains eye study. Archives of ophthalmology. 1996;114(10):1243–7.

8. Chen YY, Sheu SJ, Hu HY, Chu D, Chou P. Association between retinal vein occlusion and an increased risk of acute myocardial infarction: A nationwide population-based follow-up study. 2017;12(9):e0184016.

10. Jonas JB, Nangia V, Khare A, Sinha A, Lambat S. Prevalence and associations of retinal vein occlusions: the Central India Eye and Medical Study. Retina (Philadelphia, Pa). 2013;33(1):152–9.

11. Kolar P. Risk factors for central and branch retinal vein occlusion: a meta-analysis of published clinical data. Journal of Ophthalmology. 2014;2014:724780.

12. Fiebai B, Ejimadu CS, Komolafe RD. Incidence and risk factors for retinal vein occlusion at the University of Port Harcourt Teaching Hospital, Port Harcourt, Nigeria. Nigerian journal of clinical practice. 2014;17(4):462–6.

